# Apolipoprotein B underlies the causal relationship of circulating blood lipids with coronary heart disease

**DOI:** 10.1101/19004895

**Authors:** Tom G Richardson, Eleanor Sanderson, Tom M. Palmer, Mika Ala-Korpela, Brian A Ference, George Davey Smith, Michael V Holmes

**Author notes:** Joint senior authors (George Davey Smith, Michael V Holmes). **Correspondence to:** Tom G Richardson | MRC Integrative Epidemiology Unit (IEU), Population Health Sciences, Bristol Medical School, University of Bristol, Bristol, United Kingdom, George Davey Smith | MRC Integrative Epidemiology Unit (IEU), Population Health Sciences, Bristol Medical School, University of Bristol, Bristol, United Kingdom.

## Abstract

**Background:** Circulating blood lipids cause coronary heart disease (CHD). However, the precise way in which one or more lipoprotein lipid-related entities account for this relationship remains unclear. We sought to explore the causal relationships of blood lipid traits with risk of CHD using multivariable Mendelian randomization.

**Methods:** We conducted GWAS of circulating blood lipid traits in UK Biobank (up to n=440,546) for LDL cholesterol, triglycerides and apolipoprotein B to identify lipid-associated SNPs. Using data from CARDIoGRAMplusC4D for CHD (consisting of 60,801 cases and 123,504 controls), we performed univariable and multivariable Mendelian randomization (MR) analyses. Similar analyses were conducted for HDL cholesterol and apolipoprotein A-I.

**Findings:** GWAS identified multiple independent SNPs associated at P<5×10^−8^ for LDL cholesterol (220), apolipoprotein B (n=255), triglycerides (440), HDL cholesterol (534) and apolipoprotein AI (440). Between 56-93% of SNPs identified for each lipid trait had not been previously reported in large-scale GWAS. Almost half (46%) of these SNPs were associated at P<5×10^−8^ with more than one lipid related trait. Assessed individually using MR, each of LDL cholesterol (OR 1.66 per 1 standard deviation higher trait; 95%CI: 1.49; 1.86; P=2.4×10^−19^), triglycerides (OR 1.34; 95%CI: 1.25, 1.44; P=9.1×10^−16^) and apolipoprotein B (OR 1.73; 95%CI: 1.56, 1.91; P=1.5×10^−25^) had effect estimates consistent with a higher risk of CHD. In multivariable MR, only apolipoprotein B (OR 1.92; 95%CI: 1.31, 2.81; P=7.5×10^−4^) retained a robust effect with the estimate for LDL cholesterol (OR 0.85; 95%CI: 0.57; 1.27; P=0.44) reversing and that of triglycerides (OR 1.12; 95%CI: 1.02, 1.23; P=0.01) becoming markedly weaker.

Individual MR analyses showed a 1-SD higher HDL-C (OR 0.80; 95%CI: 0.75, 0.86; P=1.7×10^−10^) and apolipoprotein A-I (OR 0.83; 95%CI: 0.77, 0.89; P=1.0×10^−6^) to lower the risk of CHD but these effect estimates weakened to include the null on accounting for apolipoprotein B.

**Conclusions:** Apolipoprotein B is of fundamental causal relevance in the aetiology of CHD, and underlies the relationship of LDL cholesterol and triglycerides with CHD.

## Introduction

There is incontrovertible evidence that lipids play a causal role in the aetiology of coronary heart disease (CHD) [1-3]. Multiple large-scale randomized trials of lipid modifying therapies have conclusively shown that lowering of cholesterol in atherogenic lipoproteins leads to a reduction in risk of CHD[4, 5]. These findings have been recapitulated in human genetic studies using genetic variants robustly associated with low-density lipoprotein (LDL) cholesterol [6-9].

Each circulating atherogenic lipoprotein particle includes one apolipoprotein B particle but the amount of cholesterol (especially in LDL particles) and the amount of triglycerides (especially in very low-density lipoprotein particles) can vary extensively between lipoprotein particles[10]. Thus, while the concentration of LDL cholesterol and triglycerides quantifies the concentration of these lipid substances in the blood, they do not precisely quantify the number of atherogenic lipoproteins; in contrast, the concentration of apolipoprotein B particles is directly proportional to the number of circulating atherogenic particles in the blood. Evidence from human genetics supports a causal role of LDL cholesterol, triglycerides and apolipoprotein B in CHD[11-13]. While it is plausible that each lipid-related entity does individually play a causal role, it is also feasible that one trait predominates. Elucidating the comparative role of blood lipids in the aetiology of CHD has important repercussions not only in terms of a clearer understanding of the underlying pathophysiology, but also in terms of which biomarker(s) should be the focus of lipid-modifying therapeutics and might have more application in the clinical setting.

Disentangling the relationships of atherogenic lipoprotein lipids and risk of CHD is non-trivial, given the correlated nature of these traits. One such approach is to take genetic variants that associate with more than one lipid trait and scale the CHD associations for a given difference in lipid, in an attempt to identify which one or more traits appears to have a consistent effect on risk of CHD. However, such an approach makes use of indirect extrapolations and might be liable to biases, such as differential measurement error in the lipid traits. Another approach is to use Mendelian randomization (MR), a genetic approach that can facilitate an assessment of causality under certain assumptions.[14] Conventionally, MR involves the analysis of individual exposure to outcome relationships. A recently-developed extension to MR, so-called multivariable MR, permits the appraisal of multiple risk factors simultaneously. By including the genetic associations for multiple exposures in the same model, multivariable MR can elucidate which traits retain a causal relationship with an outcome of interest, through the genetic interrogation of potential confounders[15].

In this study, we sought to use human genetics to disentangle which one or more of the atherogenic lipid-related traits (apolipoprotein B, LDL cholesterol and triglycerides) is the underlying causal risk factor for CHD. We first conducted *de novo* GWAS of lipid-related traits using the UK Biobank to identify variants robustly associated with each trait. We then conducted MR analyses, including multivariable MR, to elucidate which of the atherogenic lipid traits cause CHD. Finally, we investigated whether the entity underlying the causal role of atherogenic lipid-related traits in CHD also accounted for the ‘cardioprotective’ association of HDL related phenotypes.

## Methods

### Data sources

We used data from the UK Biobank (UKBB) under application #15825 and summary estimates from CARDIoGRAMplusC4D [16]. All individual participant data used in this study were obtained from the UK Biobank study who have obtained ethics approval from the Research Ethics Committee (REC - approval number: 11/NW/0382). All participants enrolled in UK Biobank have signed consent forms.

### Data handling

Lipid-related traits in the UK Biobank were standardized/normalized using inverse rank-normalization such that the mean was 0 and standard deviation was 1, allowing comparison of effect estimates between traits.

### GWAS of lipid-related traits

We identified SNPs associated with each of the lipid-related traits using the BOLT-LMM software[17]. Analyses were adjusted for age, sex and a binary variable denoting the genotyping chip individuals were allocated to in UKBB. Population stratification and relatedness was accounted for using a mixed model after excluding individuals of non-European descent based on k-means clustering (k=4). Further details on genotyping quality control, phasing, imputation and association testing have been reported previously[18, 19]. We assigned a SNP as associated with a lipid-related trait of interest through use of a conventional GWAS threshold (P<5×10^−8^) and SNPs were binned into loci based on pair-wise LD (r^2^<0.001), with the SNP with the strongest association with the trait of interest (as defined by P value) being retained in each locus. We defined novel SNPs as those associated with the trait of interest at P<5×10^−8^ in our analyses where an association had not been previously reported at P<5×10^−8^, within 1MB and at r2<0.001, by the Global Lipids Genetics Consortium[20] (for LDL cholesterol, triglycerides and HDL cholesterol) or by Kettunen et al[21] (for apolipoprotein B or apolipoprotein A-I).

### Synthesis and characterization of genetic instruments

SNPs associating with lipid related traits at conventional GWAS thresholds (P<5×10^−8^) were taken forward to generate genetic instruments for each phenotype. We characterized the genetic instruments in several ways. First, to characterize the ‘specificity’ of individual SNPs included in each genetic instrument, we elucidated how many SNPs associated with traits other than the primary lipid trait of interest at conventional GWAS thresholds of significance (P<5×10^−8^) and used this information to generate a Venn diagram. Second, we characterize instrument ‘specificity’ by taking per-allele SNP estimates from our GWAS for each lipid trait and conducting inverse variance weighted regressions on these summary estimates to elucidate the association of genetic instruments across the various lipid related traits – these estimates are presented as standardized differences per 1-standard deviation (SD) higher genetically-predicted trait. While we recognize that this approach may be prone to inflation, the primary motivation is to characterize the associations of lipid instruments with the lipid-related traits: we do not interpret these as formal instrumental variable estimates.

### Genetic analyses to elucidate causality

We first conducted univariable Mendelian randomization (MR) analyses for each lipid-related trait. For this, we harmonized SNPs identified from our GWASs of blood lipids in UK Biobank to those SNPs available in CARDIoGRAMplusC4D by either matching the SNP directly, or by selecting proxy SNPs in high linkage disequilibrium (r2>0.8). This led to a small drop in the number of SNPs being available for MR, with a median of 93% SNPs identified in GWAS available for MR (the numbers available for each trait are provided in **Table 1**). We used the inverse variance weighted approach which, in brief takes the form of a linear regression of the SNP-outcome association regressed on the SNP-exposure association weighted by the inverse of the square of the standard error of the SNP-outcome association, with the intercept constrained at the origin. We additionally conducted univariable MR analyses using weighted median [22], weighted mode [23] and MR-Egger [24] approaches.

**Table 1.** Genetic variants identified for each trait in UK Biobank.

| **Trait** | **Number with trait measured in UK Biobank with GWAS genotyping** | **Number of SNPs identified in GWAS ($P < 5 \times 10^{-8}$)** | **Number (%) of novel\* SNPs** | **Number (%) of SNPs aligned to CARDIoGRAM-plusC4D** | **F-statistic overall and conditional** |
| --- | --- | --- | --- | --- | --- |
| **LDL cholesterol** | 440,546 | 220 | 123 (56%) | 209 (95%) | 164, 34 |
| **Triglycerides** | 441,016 | 440 | 339 (77%) | 409 (93%) | 116, 78 |
| **Apolipoprotein B** | 439,214 | 255 | 203 (80%) | 234 (92%) | 153, 36^ |
| **HDL cholesterol** | 403,943 | 534 | 383 (72%) | 490 (92%) | 124, 67 |
| **Apolipoprotein A-I** | 393,193 | 440 | 407 (93%) | 407 (93%) | 120, 62 |

\* We defined novel SNPs as those associated with the trait of interest at $P < 5 \times 10^{-8}$ where an association had not been previously reported at $P < 5 \times 10^{-8}$ , within 1MB and at $r^2 < 0.001$ by the Global Lipids Genetics Consortium[20] (for LDL cholesterol, triglycerides and HDL cholesterol) or by Kettunen et al[21] (for apolipoprotein B or apolipoprotein A-I).
^ The conditional F-statistic for apolipoprotein B when included in the multivariable MR model with LDL cholesterol and triglycerides was 36, and in the multivariable MR model that included HDL cholesterol and apolipoprotein A-I it was 66.

We next conducted multivariable MR, which is a statistical approach that allows for the association of SNPs with multiple phenotypes to be incorporated into the analysis permitting an estimation of the direct effect of each phenotype on the outcome(i.e. an effect which is not mediated by any other factor in the model[15]) see **Supplementary Figure 1** for further details. In this manuscript, we use the term ‘adjusted’ in the context of multivariable MR to mean ‘direct’ effects, i.e. the effect of a lipid trait on CHD while accounting for either mediation or confounding by another trait included in the model. For the multivariable MR analyses, we fitted a model with apolipoprotein B, LDL cholesterol and triglycerides to identify which one or more traits was responsible for the effect of “atherogenic” lipid-related traits on risk of CHD. We then took the atherogenic trait(s) that retained an effect on CHD in the multivariable MR model forward and further adjusted for apolipoprotein A-I and HDL cholesterol to assess the causal effect of HDL-related phenotypes with risk of CHD. In the setting of multivariable MR, we included all GWAS associated SNPs for all traits in the model. This meant that there were differing numbers of SNPs in the two multivariable models tested.

**Figure 1.**
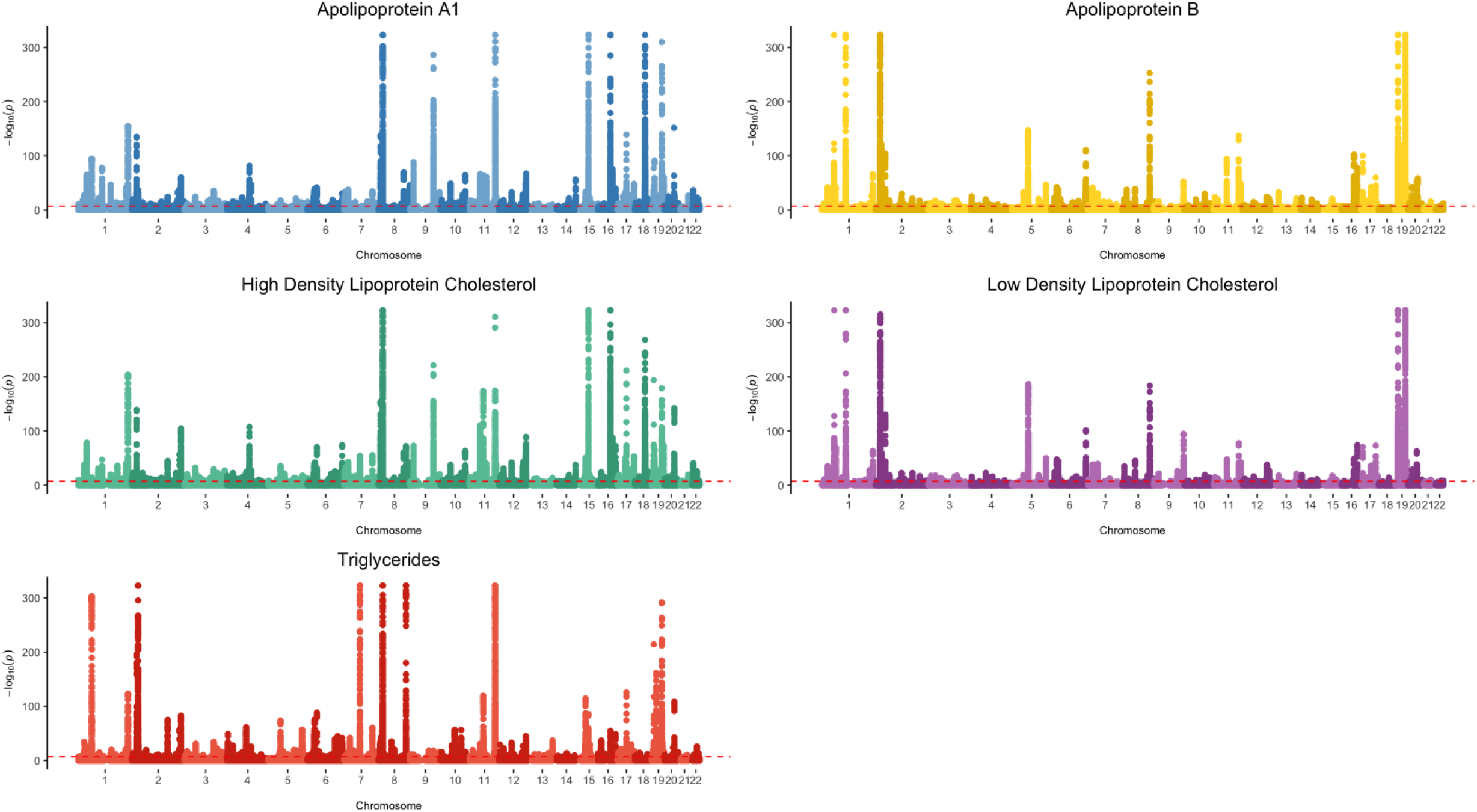
Manhattan plots showing findings from GWAS of lipid-related traits in UK Biobank.

We characterized instrument strengths in both the univariable and multivariable MR settings, as follows: for the univariable estimates, we generated the mean F-statistic, using the approximation described by Bowden et al[25]. For the multivariable estimate, we generated the conditional F-statistic [26].

### Software

The BOLT-LMM software was used to undertake GWAS [17]. MR analyses were conducted using the ‘TwoSampleMR’ R package[27]. Manhattan plots were generated using the ‘ggplot2’ package[28].The Venn diagram was generated using the online tool available at http://bioinformatics.psb.ugent.be/webtools/Venn [accessed 13th August 2019].

### Analysis

While we desisted from dichotomizing results of analyses purely on the basis of a P-value into being ‘significant’ or not[29], as a means of grading the strength of evidence against the null hypothesis, in both the univariable and multivariable Mendelian randomization analyses, we used a two-sided alpha of 0.01, on the basis of testing 5 lipid-related traits. Such a Bonferroni adjustment to account for multiple testing can be considered overly conservative given the high correlation between the lipid related traits.

## Results

### GWAS of blood lipid traits

The lipid traits were measured in 393,193 to 440,546 individuals with GWAS genotyping (**Table 1**). On GWAS, we identified a large number of independent SNPs associated at P<5×10^−8^ with each lipid related trait: 220 SNPs (of which 56% had not been previously reported) associated with LDL cholesterol, 440 (77% novel) for triglycerides, 255 (80% novel) for apolipoprotein B, 534 (72% novel) for HDL cholesterol and 440 (93% novel) for apolipoprotein A-I (**Figure 1** and **Table 1**). Full details of the SNPs associated with the lipid-related traits are provided in **Supplementary Tables 1-5**.

A considerable number (477 out of a total 1044 clumped SNPs, i.e. 46%) of SNPs used in each of the lipid-related genetic instruments showed associations at conventional GWAS significance (P<5×10^−8^) with other lipid traits (**Figure 2A**). On exploring the relationships of the genetic instruments with each lipid-related trait, we identified widespread associations (**Figure 2B**). For example, in addition to its association with apolipoprotein B, the genetic instrument for apolipoprotein B showed strong positive associations with LDL cholesterol and triglycerides and inverse associations with HDL cholesterol and apolipoprotein A-I.

**Figure 2.**
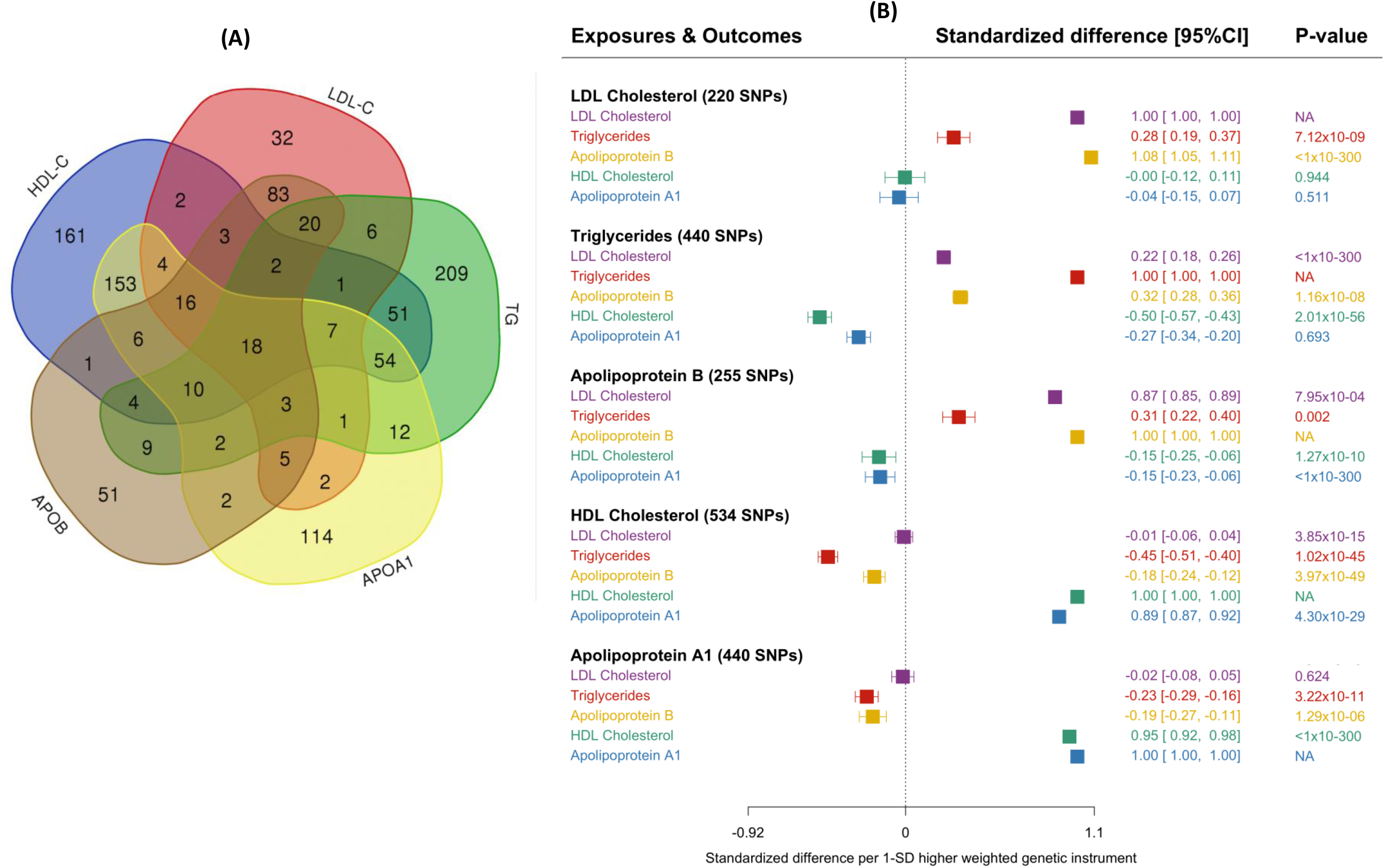
Characteristics of genetic instruments developed for lipid-related traits: (A) overlap of SNPs and (B) associations with lipids and apolipoproteins. **Legend:** In Panel A, SNPs are grouped according to whether they associate with only the primary lipid-related trait of interest, or whether they associate with other traits, based on P<5×10^−8^. Panel B displays the associations of genetic instruments with lipid related traits, using the inverse variance weighting approach. While we note the potential for overfitting of estimates displayed in Panel B, we present these data for illustrative purposes; the Mendelian randomization estimates presented in Figure 3 use a two-sample approach (with no overlapping data).

### Appraisal of LDL cholesterol, triglycerides and apolipoprotein B

On individual assessment through conventional MR, we identified LDL cholesterol, triglycerides and apolipoprotein B to have effect estimates consistent with a higher risk of CHD, using data from CARDIoGRAMplusC4D (with up to 60,801 cases) (**Figure 3A**). A 1-SD higher LDL cholesterol had an OR of 1.66 (95%CI: 1.49; 1.86; P=2.4×10^−19^) for CHD with the corresponding value for triglycerides being (OR 1.34; 95%CI: 1.25, 1.44; P=9.1×10^−16^) and apolipoprotein B (OR 1.73; 95%CI: 1.56, 1.91; P=1.5×10^−25^). Sensitivity analyses using methodological approaches that take into account potential genetic pleiotropy led to no substantive change in these estimates (**Supplementary Figure 2**).

**Figure 3.**
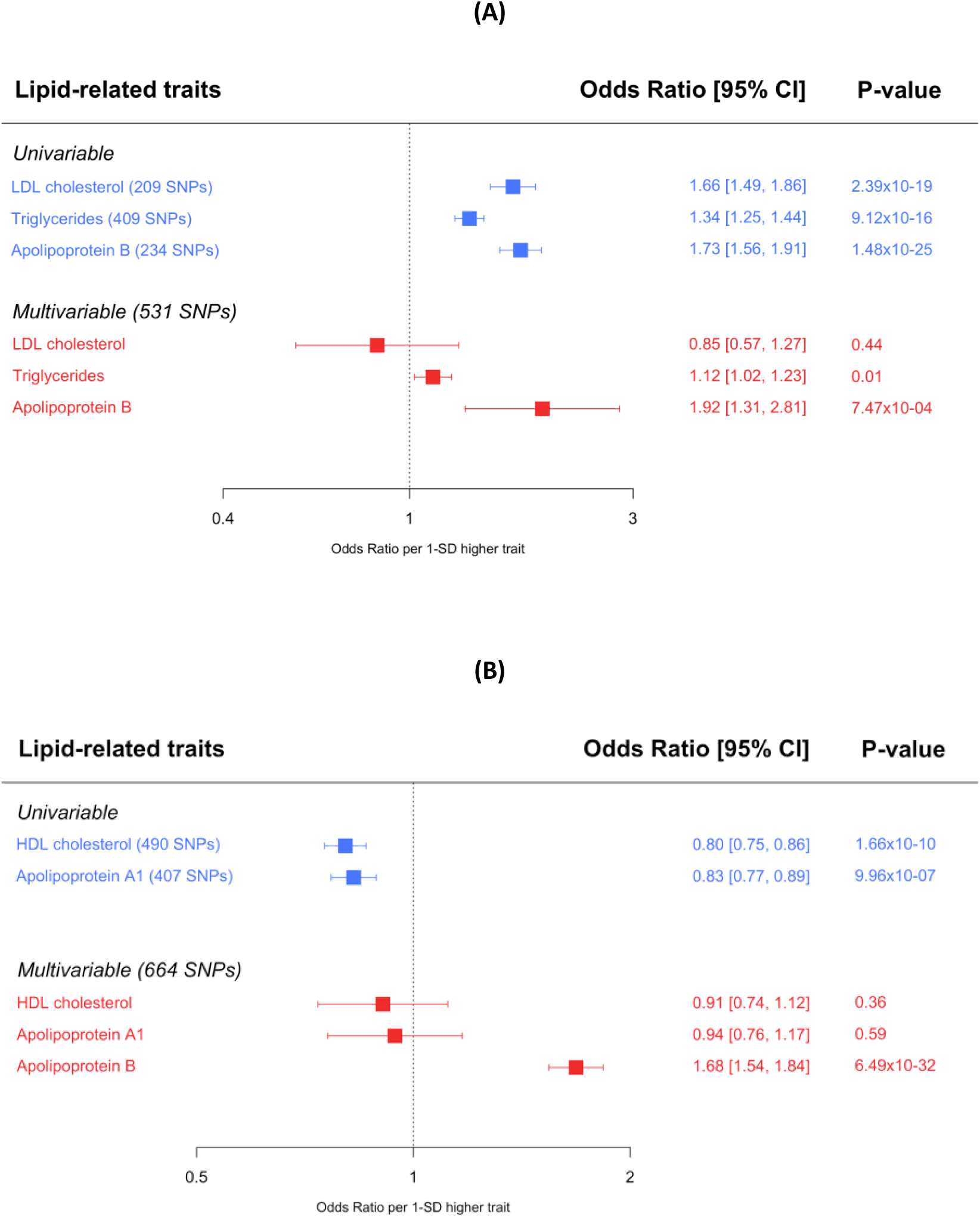
Univariable and multivariable Mendelian randomization of so-called (A) atherogenic and (B) protective lipids and apolipoproteins and risk of CHD. **Legend:** In both (A) and (B), univariable Mendelian randomization (MR) estimates were derived using the inverse variance weighted approach. For a more comprehensive repertoire of estimates derived from univariable MR approaches, please see Supplementary Figure **2**.

When LDL cholesterol, triglycerides and apolipoprotein B were assessed together in multivariable MR, only apolipoprotein B (OR 1.92; 95%CI: 1.31, 2.81; P=7.5×10^−4^) retained a robust causal relationship with CHD (**Figure 3A**). The point estimate for LDL cholesterol reversed on mutual adjustment, to yield an adjusted OR of 0.85 (95%CI: 0.57; 1.27; P=0.44). The estimate for triglycerides was weakened substantially (adjusted OR 1.12; 95%CI: 1.02, 1.23; P=0.01).

### Appraisal of HDL cholesterol and apolipoprotein A-I

Individual analysis using conventional MR showed both HDL cholesterol and apolipoprotein A-I to have effect estimates consistent with a lower risk of CHD (**Figure 3B**). The estimate for a 1-SD higher HDL cholesterol was OR 0.80 (95%CI: 0.75, 0.86; P=1.7×10^−10^) and for apolipoprotein A-I it was OR 0.83 (95%CI: 0.77, 0.89; P=9.96×10^−7^). The effect estimates for both HDL cholesterol and apolipoprotein A-I were diminished when using methodologies that are more robust to potential pleiotropy of the genetic variants used in the analysis (**Supplementary Figure 2**).

On appraisal in a multivariable MR analysis that included apolipoprotein B (which retained a causal relationship with CHD on mutual genetic adjustment for LDL cholesterol and triglycerides), the effect estimates of both HDL cholesterol and apolipoprotein A-I diminished and were not distinguishable from the null. (**Figure 3B**). The adjusted estimate for HDL-C was OR 0.91 (95%CI: 0.74, 1.12; P=0.36) and for apolipoprotein A-I it was OR 0.94 (95%CI: 0.76, 1.17; P=0.59). When adjusted for HDL cholesterol and apolipoprotein A-I, apolipoprotein B retained a robust causal effect on CHD (adjusted OR 1.68; 95%CI: 1.54, 1.84; P=6.5×10^−32^).

The F-statistics for all lipid-related genetic instruments in both the univariable and multivariable MR settings were consistent with weak instruments being an unlikely source of bias (**Table 1**).

## Discussion

Our study provides strong evidence from human genetics that supports apolipoprotein B being the underlying causal driver of the relationship of blood lipids and risk of CHD. This adds further evidence to support the hypothesis that it is the number of atherogenic lipoprotein particles, indexed by apolipoprotein B, rather than the amount of circulating cholesterol or triglycerides, that is the important driver of CHD[30]. In other words, changes in cholesterol or triglycerides that are not accompanied by commensurate changes in apolipoprotein B are unlikely to lead to altered risks of CHD.

Our GWAS identified many hundreds of variants associated with the major lipid related traits, with most SNPs identified being novel. Many SNPs identified for one lipid-related trait also showed associations with other lipid traits, highlighting their pleiotropic nature. Individual appraisal using univariable Mendelian randomization showed widespread effects of all lipid-related traits, with LDL cholesterol, triglycerides and apolipoprotein B each having effect estimates consistent with a higher risk of CHD. These findings recapitulate those reported in previous studies [8, 9, 11, 12], leading to the contemporary view that each atherogenic lipid trait might play a causal role in vascular disease. When we estimated the direct (i.e. adjusted) effect of these traits using multivariable MR (see Supplementary Figure 1 for further details), only apolipoprotein B retained a robust causal effect with CHD, with the effect of LDL cholesterol being reversed and that for triglycerides being largely diminished with only a very weak residual effect. The apparent protective associations of HDL cholesterol and apolipoprotein A-1, present on univariable MR analyses, were also markedly attenuated when direct effects conditional on apolipoprotein B were estimated. Taken together, these findings indicate that among the lipid related traits we investigated, it is apolipoprotein B, and thus the number of atherogenic lipoprotein particles, that predominates as the underlying cause of CHD.

How do these findings enhance the evidence-base relating to lipid traits and vascular disease? Large-scale observational[31], interventional[4, 5] and genetic[6-9] studies support LDL cholesterol as being causal in the aetiology of CHD. In recent years, genetic studies have provided evidence in support of triglycerides[11, 12] also playing a causal role. Both LDL cholesterol and triglycerides are carried in atherogenic lipoproteins, each containing an apolipoprotein B particle. Recent narrative reviews[32] [33] point to apolipoprotein B potentially being the necessary entity for atherosclerosis to occur, for example, through the ‘response to retention’ hypothesis, in which apolipoprotein-B containing particles become trapped in the tunica intima of the arterial wall[34]. Our study builds on recent findings[30] to provide further empirical evidence that supports this hypothesis, but our findings importantly do not discredit the causal roles that LDL cholesterol or triglycerides play in vascular disease. This is because apolipoprotein B does not occur in physiological circumstances in isolation[33], but rather is always accompanied by cholesterol and triglycerides. In light of this, our findings pinpoint that it is apolipoprotein B that is necessary in order for atherogenesis to occur. Indeed, our findings from multivariable MR are consistent with apolipoprotein B being an essential element allowing the atherogenic effects of LDL cholesterol and triglyceride to be expressed.

How do these findings aid us in the context of developing drugs that modify blood lipid concentrations and predicting their effects on risk of CHD? Drug-target Mendelian randomization studies show that, for example, modifying triglycerides through therapies such as ANGPLT3/4 inhibition may represent an emerging approach to lowering the risk of CHD[35-37] – do our findings contradict these data? Not so: our findings shed light on whether the concentrations of cholesterol and/or triglycerides that are carried by apolipoprotein B containing lipoproteins plays a role in CHD *beyond* that of apolipoprotein B. Based on these and recent data [13, 30], the primary focus of lipid modifying therapies ought to be the reduction in number of atherogenic lipoproteins (as measured by apolipoprotein B) rather than the reduction in cholesterol or triglycerides. This is especially the case where drugs have discrepant effects across these lipid traits.[10] [38] [13] Thus in predicting the vascular effects of a lipid-modifying therapeutic, apolipoprotein B can, all things being equal, be used as a reliable surrogate marker for the relative risk reduction in CHD – assuming, of course, that the drug under investigation does not display adverse events that arise either from target-mediated mechanisms, or off-target effects (notably, both can be measured directly, or extrapolated, from studies in human genetics[39]).

We note that this interpretation is in keeping with two important prior investigations that examined the concordance of CHD associations between SNPs associated with apolipoprotein B, LDL cholesterol[13] and triglycerides[30]. Indeed, one of these prior investigations conducted a form of multivariable MR analysis and obtained similar findings to those we report in the present study[30]. Importantly, the analysis that we conducted and report herein builds on these prior investigations by including the full repertoire of GWAS-associated SNPs for each of the lipid related traits (including a de novo GWAS of apolipoprotein B): such a comprehensive representation of trait-associated SNPs is necessary in order to reliably interpret the MR estimates for each of the entities included in the multivariable MR analysis.

The findings that we make have been made available by two recent advances. First, the availability of large-scale blood lipid phenotyping and GWAS genotyping in the UK Biobank, providing sufficiently large numbers to permit identification of robust genetic variants (and therefore suitable genetic instruments) in order to conduct MR of each of the lipid-related traits. Use of a single study with similar numbers of individuals with measures available for each lipid-related trait enabled GWAS and the downstream synthesis of genetic instruments for each trait in which the genetic architecture of each phenotype ought to be similarly represented, allowing for a more rigorous comparative assessment of the traits in both the univariable and multivariable MR setting. Second, methodological developments in MR to include more than one trait (so-called multivariable MR) allows for “direct” effects (i.e. the effects of an exposure on disease, taking into account potential confounding and mediation by other traits) of multiple exposures to be assessed simultaneously and without the risk that this introduces forms of bias (such as collider bias)[15]. It is this methodological approach that allows the deduction that we make: that apolipoprotein B underlies the causal effects of lipid-related traits with risk of CHD. We note here an important theme that emerges: the discrepancy between our findings and those derived from other MR approaches that hitherto have been used in contemporary MR studies (reflected by the univariable MR estimates we present in Supplementary Figure 2). While other approaches such as MR-Egger and weighted median MR can provide reliable tests of causation even in the presence of confounding through unbalanced horizontal pleiotropy[40] (as evidenced by the diminution of the HDL cholesterol association with risk of CHD on MR-Egger, Supplementary Figure 2), such approaches notably do not, with a few exceptions [41, 42], allow simultaneous statistical adjustment for multiple traits. The repertoire of univariable MR analyses that seek to act as sensitivity analyses for potential pleiotropy each makes a different series of assumptions [22]. In the context that genetic confounding affects the majority of SNPs used in the genetic instruments, and when such confounding is present in a dose-response manner (i.e. on average, SNPs that increase the exposure of interest also increase the confounder of interest), this violates the ‘inSIDE’ assumption[24] and the MR analyses yield biased estimates. This is why the MR estimates for LDL cholesterol and triglycerides remain seemingly robust to MR Egger and weighted median MR approaches. In this context, multivariable MR analysis can help when the traits included in the analysis account fully for the unbalanced, dose-related, horizontal pleiotropy. In the scenario that we investigate, apolipoprotein B does just so, permitting us to conclude that it is apolipoprotein B that is ultimately responsible for the underlying causal relationship of blood lipids and risk of CHD.

In conclusion, our findings demonstrate that apolipoprotein B underlies the causal effect of lipids on CHD and that it is the trait that is responsible for the associations of LDL cholesterol, triglycerides, HDL cholesterol and apolipoprotein A-I with the risk of CHD.

## Data Availability

GWAS data will become available once the manuscript is In Press

## Acknowledgements

We are immensely grateful to study participants of the UK Biobank.

